# Food Insecurity, Diet Quality and Associated Factors among Mothers and Children in Rural North-East Ghana

**DOI:** 10.1101/2025.11.17.25340374

**Authors:** Vincent Adocta Awuuh, Charles Apprey, Reginald Adjetey Annan

**Affiliations:** Department of Biochemistry and Biotechnology, Kwame Nkrumah University of Science and Technology, Private Mail Bag, Ghana; Department of Nutrition and Dietetics, College of Nursing and Midwifery, Nalerigu, Ministry of Health, P. O. Box 10, Nalerigu, Ghana

**Keywords:** *Food insecurity*, *dietary diversity*, *maternal nutrition*, *child nutrition*, *household*, *malnutrition*, *correlates*

## Abstract

**Introduction:** Food insecurity, poor diet quality, and malnutrition remain major public health concerns in sub-Saharan Africa. Despite high nutritional vulnerability in North-East Ghana, limited research has comprehensively examined relationships between food insecurity, diet quality, and maternal-child nutrition outcomes. This study addresses critical gaps by providing an integrated analysis of these factors among mother-child dyads in rural North-East Ghana.

**Methodology:** A cross-sectional study was conducted among 248 households in East Mamprusi Municipality [June–July 2023]. Food security was assessed using the Household Food Insecurity Access Scale [HFIAS] and diet quality using the Diet Quality Questionnaire [DQQ]. Data were analysed using chi-square, Spearman’s correlation, and Mann–Whitney U tests to identify determinants of dietary diversity.

**Results:** Most households [81.5%] were food insecure. Health insurance coverage [χ² = 4.31, p = 0.038] and partner occupation [χ² = 13.09, p = 0.023] were key correlates of food security. Women’s dietary diversity was moderate [mean score = 4.45], with 29.8% consuming all five essential food groups. Vegetable consumption [76.2%] exceeded fruit intake [46.0%]. Among children, 97.2% were breastfed; however, only 44.4% met the minimum dietary diversity, and 27.2% met the minimum acceptable diet. Monthly food expenditure showed the strongest association with maternal [r = -0.44, p < 0.001] and child [r = -0.397, p < 0.001] dietary diversity. Polygamous households had poorer dietary outcomes for both mothers [p = 0.007] and children [p = 0.003]. Food insecurity was negatively correlated with maternal haemoglobin [ρ = -0.283, p < 0.001], child height-for-age [ρ = -0.166, p = 0.009], and weight-for-height [ρ = -0.158, p = 0.013].

**Conclusion:** Food insecurity and suboptimal diet quality persist in rural Ghana, influenced by factors such as economic status, household structure, and health service coverage. Place-based, co-designed interventions addressing economic empowerment, health coverage, and nutrition empowerment that target emerging unhealthy dietary patterns are vital for improving maternal and child nutrition and health outcomes.

## Introduction

Food insecurity and inadequate diet quality are pressing global health concerns [1], disproportionately affecting vulnerable populations such as women and children [2]. Across low-and middle-income countries, limited access to safe and nutritious foods contributes to high rates of malnutrition, micronutrient deficiencies, and poor health outcomes [3, 5]. In 2019, nearly 690 million people were undernourished, with numbers rising steadily since 2014 [5]. The COVID-19 pandemic further exacerbated the situation, potentially adding 83–132 million more undernourished individuals [5]. By 2021, an estimated 828 million people were affected by hunger, a staggering increase of 150 million since 2019, while severe food insecurity impacted 924 million people in the same year [6]. Undernutrition contributes to 45% of preventable deaths among children under five years in low- and middle-income countries [7]. These trends highlight the urgent need to accelerate progress toward Sustainable Development Goal 2 [Zero Hunger] [5].

Sub-Saharan Africa bears a disproportionate share of the global food insecurity burden [8]. Approximately 239 million people in the region suffered from hunger, with food insecurity worsening [9]. Conflict, economic slowdowns, climate change, and the pandemic continue to undermine maternal and child nutrition in the region[10]. Additional factors exacerbating food insecurity include weak economic growth, gender inequality, corruption, and inadequate investment in agricultural research and infrastructure [9]. Ghana has made significant progress in reducing poverty and child undernutrition, achieving Millennium Development Goal targets on extreme poverty and hunger [11]. However, substantial regional disparities persist, particularly between northern and southern regions, with large rural-urban gaps remaining across most dimensions of food and nutrition security [11]. Child malnutrition disproportionately affects the poor, with socioeconomic inequality primarily associated with poverty, healthcare access, and regional disparities [12]. Currently, approximately 1.5 million Ghanaians are estimated to be food insecure, while undernutrition, overnutrition, and micronutrient deficiencies persist across all life stages [13]. Key challenges include poverty, climate change, rapid urbanisation, population growth, gender inequalities, and poor infrastructure [13]. Ghana increasingly faces a double burden of malnutrition, with rising overweight and obesity rates alongside persistent undernutrition, creating new public health challenges [11]. Northern Ghana is identified as among the country’s most vulnerable regions to climate variability, with the area historically being the poorest [14]. The prevalence of moderate and severe household hunger in Northern Ghana has been reported at 25.9% and 6.8%, respectively [15]. Similarly, in the East Mamprusi Municipality, 71.9% of households experience food insecurity, which has been linked to maternal mental health challenges [16].

The North-East Region, where the East Mamprusi Municipality is located, is among the poorest and most vulnerable regions in Ghana. The region’s semi-arid climate, short and erratic rainy season, and frequent droughts and floods severely undermine agricultural productivity [17]. Its geographic isolation, poor road networks, and limited infrastructure restrict access to diverse foods and markets, driving higher food prices compared with Southern Ghana [18]. More than 70% of the population lives below the poverty line, with livelihoods heavily dependent on subsistence farming and traditional crops such as millet, sorghum, and groundnuts. These structural, geographic, and cultural conditions make the region particularly prone to seasonal hunger and chronic food insecurity, yet nutrition research in this region remains limited [19].

Despite the region’s high nutrition vulnerability, few studies have comprehensively examined the complex relationships between food insecurity, diet quality, and maternal–child nutrition outcomes in the North-East Region. Existing research in Ghana has largely focused on national or zonal averages, masking important local variations, or has examined isolated outcomes such as child stunting or maternal anaemia [16]. Seasonal fluctuations in food insecurity, which are especially pronounced in this region due to its single rainy season, are also poorly documented in relation to maternal and child diet quality. Without significant action, food security and child nutrition in Sub-Saharan Africa are expected to deteriorate further [20]. This study addresses these critical gaps by providing the first integrated analysis of food insecurity, diet quality, and their correlates among mother–child dyads in rural North-East Ghana. The study carries strong policy relevance, as its findings are expected to inform targeted nutrition interventions

## Materials and Methods

### Ethics Statement

This research received ethical clearance from the Committee on Human Research, Publication and Ethics at Kwame Nkrumah University of Science and Technology, Kumasi **(approval number: CHRPE/AP/149/23),** and authorisation was granted by the Ghana Health Service’s East Mamprusi Municipal Health Directorate **(reference: GHS/NER/EMM/10/2023).** The trial was formally registered under the Pan African Clinical Trial Registry with registration number **PACTR202412697952932.** All study participants provided written informed consent before enrollment, and parental/guardian consent was secured for minor participants. Involvement in the study was completely voluntary, free from any form of pressure, and participants retained the right to discontinue participation at any point without facing any repercussions. All data collected during the study were kept strictly confidential. Before commencing data collection, appropriate community engagement protocols were followed, including consultation sessions with local traditional authorities and community leaders to ensure cultural appropriateness and respectful conduct throughout the research process.

### Study Area

The study was conducted in the East Mamprusi Municipality, located in the North East Region of Ghana. The area is characterised by high levels of food insecurity, with a significant proportion of households engaged in subsistence farming. The region experiences an unimodal rainfall pattern, with the dry season lasting from November to April, influencing food availability and dietary patterns [21].

### Study Design and Participants

A cross-sectional study design assessed household food insecurity and dietary adequacy among women aged 18-49 and children aged 6-59 months. The study included households with children in the specified age group and their mothers or primary caregivers.

### Sample Size and Recruitment

Sample size was determined using G*Power version 3.1.9.7 for a two-tailed correlation analysis, with α = 0.05 and 80% power. A medium effect size [r = 0.30] was specified, based on prior nutrition and public health studies that have reported moderate associations between household food insecurity and diet quality [22;23]. Based on these assumptions, a minimum sample of 85 households was required. To accommodate potential incomplete responses, missing data, and to enhance statistical power for subgroup analyses, recruitment was extended to 248 households. This final sample size exceeded the minimum requirement by approximately 190%, ensuring adequate power. Participants were recruited using a multistage sampling approach. First, communities within the East Mamprusi Municipality were stratified by geographic location. Next, households were randomly selected from each stratum proportionate to population size, ensuring fair representation across different areas. Finally, within each selected household, mothers with at least one child were identified and enrolled. This sampling approach was adopted to ensure diversity in the sample while minimising selection bias.

### Inclusion and Exclusion Criteria

Households with at least one woman of reproductive age [18–49 years] and a child aged 6–59 months were eligible for inclusion. This group forms part of the vulnerable group for malnutrition. Households where the eligible participants were suffering from fever or diarrhoea at the time of data collection were excluded. The justification was to prevent bias, as illness may temporarily alter appetite, food intake, or feeding practices, leading to inaccurate assessments of usual diet quality and food security. Households that declined consent were also excluded.

### Data Collection

Data collection involved structured household surveys and nutritional assessments. Trained field staff conducted face-to-face interviews using a pretested questionnaire, mounted on KOBO Collect. The Household Food Insecurity Access Scale [HFIAS] tool was used to assess food security levels. The Diet Quality Questionnaire [DQQ] was employed to assess dietary adequacy among mother-child pairs. Anthropometric data, including weight, height, age, and blood samples of mothers and children, were collected and analysed for key nutritional indicators. To ensure accuracy and consistency, field enumerators conducted duplicate measurements of participants to minimise measurement error.

### Food Security Assessment

Food security was assessed using the Household Food Insecurity Access Scale [HFIAS] developed by USAID’s FANTA project, adapted for the study population [24]. The HFIAS is a validated instrument containing 9 occurrence questions with corresponding frequency questions, designed to assess food insecurity experiences over the preceding 30 days across three distinct domains: Anxiety and uncertainty about food supply [capturing worry about household food access], insufficient food quality [including dietary variety and food preferences] and insufficient food quantity [measuring inadequate food intake and hunger] [25]

### HFIAS Scoring Protocol

The HFIAS scoring followed standardised procedures with systematic coding. Each of the 9 occurrence questions was coded as 0 [condition did not occur] or 1 [condition occurred]. When occurrence equalled 1, frequency was coded as: 1 = Rarely [1-2 times in the past 30 days], 2 = Sometimes [3-10 times in the past 30 days], or 3 = Often [more than 10 times in the past 30 days]. The total HFIAS score was calculated by summing all frequency responses, ranging from 0-27, with higher scores indicating greater severity of food insecurity [24].

### Food Security Classification

Household food security was classified according to the Household Food Insecurity Access Prevalence [HFIAP] indicator [24]. Although the original HFIAP protocol categorises households into four groups [Food Secure, Mild, Moderate, and Severe], for statistical analysis in this study, categories 2, 3 & 4 for Mild, Moderate, and Severe, respectively, were collapsed into a single food insecure group. This ensured adequate cell counts for Chi-square tests while maintaining analytical clarity.

**Category 1** – Food secure constituted households experiencing no food insecurity conditions or only occasional worry about food access, with no compromises to food quality or quantity.

*Classification criteria*:[[Q1a=0 or Q1a=1] and Q2=0 and Q3=0 and Q4=0 and Q5=0 and Q6=0 and Q7=0 and Q8=0 and Q9=0].

**Category 2** – Food Insecure [Mild, Moderate, and Severe] comprises households experiencing any degree of food insecurity, ranging from anxiety about food supply and reduced dietary variety to meal size reductions, hunger, or full-day fasting.

*Classification criteria:* From *Mild Food Insecurity*: [Q1a=2 or Q1a=3 or Q2a=1 or Q2a=2 or Q2a=3 or Q3a=1 or Q4a=1] and Q5=0 and Q6=0 and Q7=0 and Q8=0 and Q9=0

OR From *Moderate Food Insecurity:* [Q3a=2 or Q3a=3 or Q4a=2 or Q4a=3 or Q5a=1 or Q5a=2 or Q6a=1 or Q6a=2] and Q7=0 and Q8=0 and Q9=0

OR From *Severe Food Insecurity*: Q5a=3 or Q6a=3 or Q7a=1 or Q7a=2 or Q7a=3 or Q8a=1 or Q8a=2 or Q8a=3 or Q9a=1 or Q9a=2 or Q9a=3

This approach preserved the original HFIAS scoring procedure to determine individual household food insecurity status before collapsing the categories for analysis.

### Diet Quality Assessment

The diet quality of women and children was assessed using the Diet Quality Questionnaire [DQQ], a validated tool for population-level monitoring of dietary patterns and non-communicable disease [NCD]-related risk and protective behaviours [26]. The DQQ relies on a standardised 24-hour recall that captures food group consumption across 29 categories. For this study, indicators were calculated separately for women of reproductive age and children aged 6–59 months. Data were collected using the DQQ Indicator Calculator, which processes responses in a standardised CSV template and uploads them to the tool calculator to automatically compute dietary quality indicators.

### DQQ Indicators for Women

For women, several dietary adequacy and quality indicators were computed. The Minimum Dietary Diversity for Women [MDD-W] assessed whether participants consumed at least 4-6 food groups for children and ≥5 for women of reproductive age out of the recommended food groups in the previous 24 hours [27;28]. The ALL-5 indicator measured whether all five essential food groups, starchy staples, vegetables, fruits, pulses/nuts/seeds, and animal-source foods were consumed. Additional binary indicators captured the consumption of specific food items, including at least one vegetable, one fruit, one pulse, nut, or seed, one animal-source food, and one starchy staple.

Dietary risk and protective behaviours were also measured. The NCD-Protect Score [0–9] reflected the intake of health-promoting foods such as whole grains, pulses, nuts, vitamin A-rich vegetables, dark green leafy vegetables, and fruits. In contrast, the NCD-Risk Score [0–9] captured the consumption of foods associated with increased NCD risk, including processed meats, sugar-sweetened beverages, sweet foods, salty or fried snacks, and deep-fried foods. From these, the Global Dietary Recommendations [GDR] Score [0–18] was calculated [NCD-Protect – NCD-Risk + 9], with higher scores indicating better adherence to WHO dietary guidelines [26].

### DQQ Indicators for Children

For children aged 6–59 months, indicators focused on infant and young child feeding practices recommended by the World Health Organisation [WHO]. These included ever breastfed, exclusive breastfeeding during the first two days of life, and continued breastfeeding between 12–23 months. For non-breastfed children, the minimum milk feeding frequency was assessed. Additional indicators captured minimum dietary diversity, minimum meal frequency, and the minimum acceptable diet among children aged 6–23 months.

Dietary quality was further assessed using binary indicators of food group consumption. These included egg and/or flesh food consumption, whole grain consumption, and processed meat consumption. Indicators of unhealthy dietary patterns included sweet beverage consumption, sweet food consumption, savoury and fried snack consumption, and zero fruit or vegetable consumption. Bottle feeding practices among children aged 0–23 months were also documented.

### Socio-Demographic and Nutritional Data Collection

Socio-demographic data were collected using a structured questionnaire administered to the primary caregivers of households. Information captured included household size, maternal age, educational level, marital status, occupation, and household income sources and food expenditure. Additional household characteristics, such as access to water, sanitation, and electricity, were also documented to provide contextual factors that may influence food security and diet quality.

Anthropometric measurements and blood samples were taken for both women of reproductive age and children aged 6–59 months following standardised WHO procedures. For women, weight was measured using a calibrated digital scale to the nearest 0.1 kg, while height was measured using a portable stadiometer to the nearest 0.1 cm. For children, weight was measured using a UNICEF electronic infant scale and the mother–child tared weighing method [for younger children]. Recumbent length [for children under 24 months] or standing height [for those 24 months and older] was measured with a length/height board.

All anthropometric measurements were taken twice and averaged to improve accuracy. Enumerators were trained before data collection to ensure consistency and reliability. The anthropometric indicators generated included body mass index [BMI] for women, and weight-for-age [WAZ], height-for-age [HAZ], and weight-for-height [WHZ] classifications for children, which were compared against WHO growth standards to assess nutritional status. A full blood count was performed on blood samples to determine the child’s/mother’s haemoglobin levels for the detection of anaemia status.

### Statistical Analysis

Data were analysed using descriptive statistics [frequencies, percentages, and means] to summarize socio-demographic characteristics, household food security status, and diet quality indicators. Chi-square [χ²] tests were performed to assess associations between household food security and socio-demographic characteristics.

Spearman’s rank correlation analysis was employed due to the non-parametric nature of the data [Kolmogorov-Smirnov test, p < 0.001; Shapiro-Wilk test, p < 0.001] to examine the relationships between the Household Food Insecurity Access Score [HFIAS] and various nutritional outcomes. Mann-Whitney U tests were conducted to identify the determinants of maternal dietary diversity achievement [meeting the Minimum Dietary Diversity for Women - MDDW] and child dietary diversity achievement [meeting the Minimum Dietary Diversity - MDD]. These non-parametric tests compared median values between groups that did and did not achieve dietary diversity targets. Effect sizes [r] were calculated to determine the practical significance of observed differences, with values of 0.1, 0.3, and 0.5 representing small, medium, and large effect sizes, respectively. Statistical significance was set at p < 0.05 for all analyses. Data analysis was performed using SPSS.

## Results

### Socio-Demographics of Study Population

Table 1 presents the socio-demographic characteristics of the study population. Almost all respondents were married [97.6%], with approximately half having one wife [51.2%] and the remainder having multiple wives [48.8%]. The majority were either Christian [52.4%] or Muslim [46.8%]. Mamprusi [56.5%] and Komkomba [27.4%] were the dominant ethnic groups.

**Table 1:** Socio-Demographic Characteristics of Study Participants

| Variable | Category | Frequency [n] | Percent [%] |
| --- | --- | --- | --- |
| Marital Status | Married | 242 | 97.6 |
|  | Unmarried/Divorced | 6 | 2.4 |
| Number of Wives | One wife | 127 | 51.2 |
|  | More than one wife | 121 | 48.8 |
| Religion | Christian | 130 | 52.4 |
|  | Muslim | 116 | 46.8 |
|  | Others | 2 | 0.8 |
| Ethnicity | Mamprusi | 140 | 56.5 |
|  | Komkomba [Likpakpa] | 68 | 27.4 |
|  | Others | 40 | 16.1 |
| Mother's Education | No formal education | 209 | 84.3 |
|  | Middle/JSS/JHS | 31 | 12.5 |
|  | Secondary/Technical/Vocational | 8 | 3.2 |
| Mother's Occupation | Farmer | 182 | 73.4 |
|  | Housewife | 38 | 15.3 |
|  | Others | 28 | 11.3 |
| Partner's Occupation | Farmer | 216 | 87.1 |
|  | Government employee | 8 | 3.2 |
|  | Others | 24 | 9.7 |
| Household Head Gender | Male | 245 | 98.8 |
|  | Female | 3 | 1.2 |
| Type of Household | Nuclear | 118 | 47.6 |
|  | Extended | 130 | 52.4 |
| Child Gender | Male | 139 | 56.0 |
|  | Female | 109 | 44.0 |

*\*\*Mean age of children: 20.9 [ $\pm 11.4$ ] months; range = 6.6–55.8 months. \*\* \*\*\*Mean age of mothers: 29.3 [ $\pm 7.1$ ] years; range = 16–47 years\*\*\**

Most mothers had no formal education [84.3%] and were primarily engaged in farming [73.4%], while their partners were predominantly farmers [87.1%]. Nearly all households were male-headed [98.8%], and household types were almost evenly divided between nuclear [47.6%] and extended [52.4%].

The mean age of children was 20.9 [±11.4] months, with a slightly higher proportion of males [56.0%] compared to females [44.0%].

### Food Insecurity Prevalence

Out of the total 248 households that participated in this study, the majority [81.5%, n=202] were classified as food insecure, while only 18.5% [n=46] achieved a food security status [Table 2].

**Table 2.**
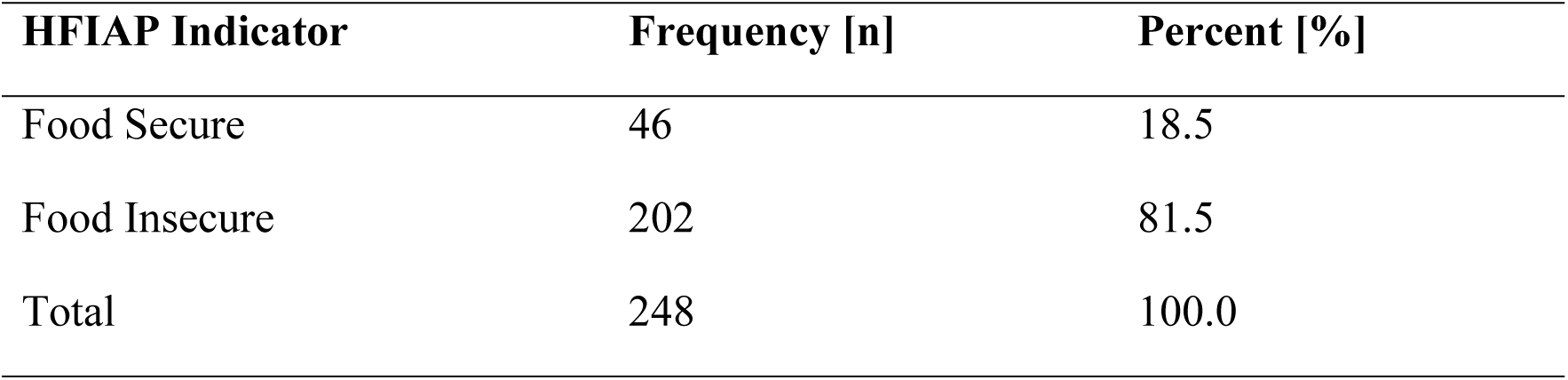
Household Food Insecurity Prevalence

| HFIAP Indicator | Frequency [n] | Percent [%] |
| --- | --- | --- |
| Food Secure | 46 | 18.5 |
| Food Insecure | 202 | 81.5 |
| Total | 248 | 100.0 |

### Sociodemographic Correlates of Household Food Insecurity

Analysis of sociodemographic factors revealed that health insurance coverage was significantly associated with household food security [χ² = 4.31, df = 1, p = 0.038]. Households without active health insurance demonstrated substantially higher rates of food insecurity [84.4%] compared to those with insurance coverage [72.6%]. Partner occupation also showed a significant association with food security status [χ² = 13.092, p = 0.023], with traders exhibiting the lowest food insecurity rates [37.5%] compared to farmers [82.4%] and other occupational categories [Table 3].

**Table 3.**
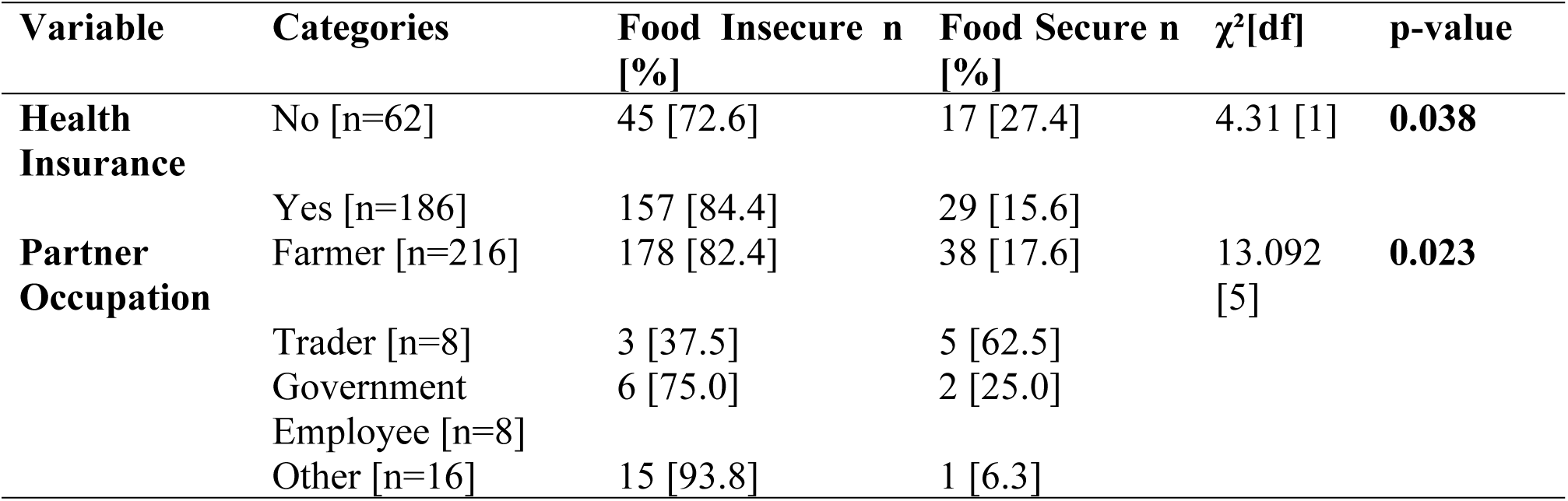
Significant Sociodemographic Associations of Household Food Insecurity

### Food Insecurity and Nutritional Outcomes

Spearman’s rank correlation analysis revealed significant negative associations between household food insecurity [HFIAS] and key nutritional indicators [Table 4]. The strongest correlation was observed with maternal hemoglobin levels [ρ=-0.283, p<0.001], indicating that higher food insecurity was associated with lower maternal hemoglobin status. Food insecurity was also significantly correlated with reduced mother-child monthly food expenditure [ρ=-0.154, p=0.015], lower overall Essential Nutrition Actions practiced [ENAs] [ρ=-0.213, p=0.001], and poorer child anthropometric outcomes including height-for-age Z-score [ρ=-0.166, p=0.009] and weight-for-height Z-score [ρ=-0.158, p=0.013].

**Table 4.**
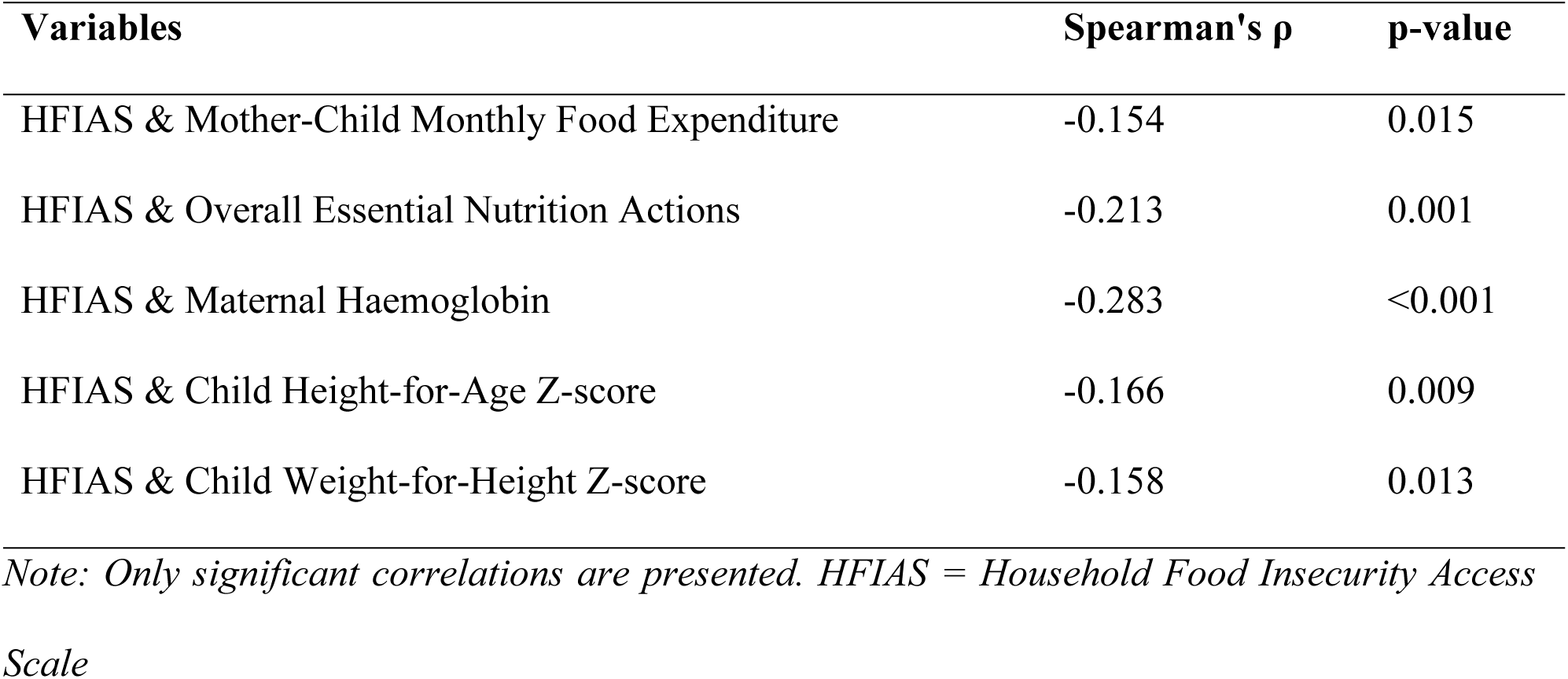
Correlations Between Food Insecurity and Nutritional Outcomes

### Maternal Diet Quality Achievement and Determinants

Only 29.8% of women met the “All-5” protective food group criteria, consuming all five essential food groups. The mean of the minimum dietary diversity score for women [MDDW] was 4.45 out of 10 possible food groups. Analysis of dietary patterns revealed that 76.2% consumed at least one vegetable daily, while only 46.0% consumed at least one fruit. Animal-source food consumption was achieved by 61.3% of women, and 83.1% consumed starchy staples as their primary energy source. Regarding composite diet quality indicators, women achieved a mean NCD-Protect score of 3.67 out of 9 possible points, indicating moderate consumption of protective foods against non-communicable diseases. The mean NCD-Risk score was 0.70 out of 9, suggesting relatively low consumption of foods associated with increased disease risk. Overall, the Global Diet Quality Score averaged 11.97 out of 18 possible points, reflecting moderate overall diet quality among the study population [Table 5].

**Table 5.** Women’s Diet Quality Indicators and Scores

| Indicator | Percentage/Mean Score |
| --- | --- |
| **Core Dietary Diversity Indicators** |  |
| All 5 food groups [out of 5] | 29.8% |
| At least one vegetable | 76.2% |
| At least one fruit | 46.0% |
| At least one pulse, nut or seed | 69.0% |
| At least one animal-source food | 61.3% |
| At least one starchy staple | 83.1% |
| Dietary Diversity Score [DDS]- *mean* * | 4.45* |
| **Specific Food Group Consumption** |  |
| Zero vegetable or fruit consumption | 20.2% |
| At least one vegetable or fruit | 79.8% |
| Pulse consumption | 39.1% |
| Nuts or seeds consumption | 60.1% |
| Whole grain consumption | 76.2% |
| At least one whole grain, pulse, nut or seed | 85.5% |
| **Processed and Ultra-processed Foods** |  |
| Processed meat consumption | 0.8% |
| Salty or fried snack consumption | 10.9% |
| Deep-fried food | 4.4% |
| Sweet food consumption | 22.2% |
| Soft drinks [sodas, energy drinks, sports drinks] | 13.7% |
| Fast food or instant noodles | 6.5% |
| More than one sugary food or beverage | 21.8% |
| More than one salty ultra-processed food | 3.6% |
| **Composite Diet Quality Scores** |  |
| NCD-Protect score [out of 9]- *mean* * | 3.67* |
| NCD-Risk score [out of 9]- *mean* * | 0.70* |
| Global Diet Quality Score [out of 18]- *mean* * | 11.97* |
| **Diet Pattern Classifications** |  |
| Protective Food Consumption [PFC] | 40.3% |
| Unhealthy Food Consumption [UFC] | 21.8% |
| Healthy Diet Pattern | 23.4% |

*Note: Values are presented as percentages unless otherwise specified for mean scores [\*]. NCD = non-communicable disease; PFC = Protective Food Consumption; UFC = Unhealthy Food Consumption*

Mann-Whitney U analysis identified household polygamy and monthly food expenditure as the primary determinants of maternal dietary diversity achievement [Table 6]. Women in monogamous households showed a trend toward better MDDW achievement compared to those in polygamous households, though this difference approached but did not reach statistical significance [p=0.086]. Monthly mother-child food expenditure demonstrated the strongest association with maternal dietary diversity [Z=-6.558, p<0.001, r=-0.44, large effect size], with women meeting MDDW having consistently higher median food expenditures.

**Table 6.** Determinants of Maternal Dietary Diversity Achievement

| Variable | MDDW Status | N | Median [IQR] | Z | p-value | Effect Size [r] |
| --- | --- | --- | --- | --- | --- | --- |
| Number of Wives | Did **NOT** meet | 175 | 2 [1-2] | -2.706 | 0.007 | -0.17 |
|  | Met | 72 | 1 [1-2] |  |  |  |
| Working-age Members | Did **NOT** meet | 176 | 4 [2-7] | -3.003 | 0.003 | -0.19 |
|  | Met | 72 | 5 [4-7] |  |  |  |
| Monthly Food Expenditure | Did **NOT** meet | 176 | 240[120-260]<br>GHS | -6.558 | <0.001 | -0.44 |
|  | Met | 72 | 240[240-260]<br>GHS |  |  |  |

*GHS = Ghana Cedis, MDDW – Minimum Dietary Diversity for Women, N – The number of observations or participants included in the analysis, IQR [Interquartile Range], Z – The Z-value [Z-statistic]*

### Child Dietary Quality and Associated Factors

Child minimum dietary diversity [MDD] was achieved by 44.4% of children, while only 27.2% met the minimum acceptable diet criteria [combining both dietary diversity and meal frequency]. Breastfeeding practices were generally favourable, with 97.2% of children having been breastfed and 75.6% exclusively breastfed during the first two days after birth [Table 7].

**Table 7.** Child Feeding Practices and Dietary Quality Indicators

| Indicator | Percentage |
| --- | --- |
| **Breastfeeding Practices** |  |
| Ever breastfed | 97.2% |
| Exclusively breastfed for the first two days after birth | 75.6% |
| Continued breastfeeding for 12-23 months | 90.6% |
| Bottle feeding [0-23 months] | 32.4% |
| **Minimum Feeding Requirements** |  |
| Minimum dietary diversity [6-23 months] | 44.4% |
| Minimum meal frequency [6-23 months] | 51.5% |
| Minimum milk feeding frequency for non-breastfed children [6-23 months] | 66.6% |
| Minimum acceptable diet [6-23 months] | 27.2% |
| All-5 food groups [6-23 months] | 13.8% |
| **Specific Food Consumption** |  |
| Egg and/or flesh food consumption [6-23 months] | 33.3% |
| Whole grain consumption [6-23 months] | 50.0% |
| Zero vegetable or fruit consumption [6-23 months] | 22.2% |
| **Unhealthy Food Consumption** |  |
| Sweet beverage consumption [6-23 months] | 41.6% |
| Sweet food consumption [6-23 months] | 25.0% |
| Savoury and fried snack consumption [6-23 months] | 11.1% |
| Processed meat consumption [6-23 months] | 5.5% |
| Overall unhealthy food consumption [6-23 months] | 25.0% |

Mann-Whitney U analysis confirmed household structure and food expenditure as key determinants of child dietary diversity [Table 8]. Children who achieved MDD came from households with significantly fewer wives [median = 1.0 vs. 2.0, Z = -2.946, p = 0.003, r = -0.187]. The mother-child monthly food expenditure showed the strongest association with child MDD achievement [Z = -6.260, p < 0.001, r = -0.397, large effect size]. Additionally, household food insecurity scores were significantly lower among children who achieved MDD [Z=-2.749, p=0.006, r=-0.174], indicating better food security conditions in households with adequate child dietary diversity.

**Table 8.** Determinants of Child Dietary Diversity Achievement

| Variable | MDD<br>Status | N | Median [IQR] | Z | p-value | Effect<br>Size [r] |
| --- | --- | --- | --- | --- | --- | --- |
| Number of Wives | **Not** Met | 139 | 2.0 [1-2] | -2.946 | 0.003 | -0.187 |
|  | Met | 108 | 1.0 [1-2] |  |  |  |
| Monthly Food<br>Expenditure | **Not** Met | 140 | 240.0 [120-260] GHS | -6.260 | <0.001 | -0.397 |
|  | Met | 108 | 240.0 [240-260] GHS |  |  |  |
| HFIAS Score | **Not** Met | 140 | 12.0 [6-19] | -2.749 | 0.006 | -0.174 |
|  | Met | 108 | 12.0 [6-19] |  |  |  |

*HFIAS = Household Food Insecurity Access Scale; GHS = Ghana Cedis*

## Discussion

This study provides insights into household food security, maternal and child dietary quality, and the correlates of these in the East Mamprusi Municipality of northern Ghana. The findings highlight the persistence of severe food insecurity in rural Ghana, where over four-fifths of households [81.5%] were food-insecure, with only a small minority [18.5%] achieving food security. This rate is higher than the 71.9% reported in similar studies [16], in the same municipality, who employed the same sampling method but used the Food Insecurity Experience Scale [FIES] instead of the Household Food Insecurity Access Scale [HFIAS] and collected data in September–October 2020, rather than June–July 2023, as in this study, suggesting worsening food security conditions. The most difficult time in terms of food insecurity in northern Ghana is the months of June and July [29].

These findings underscore the persistent vulnerability of rural northern Ghana to chronic food insecurity. Research across sub-Saharan Africa reveals widespread food insecurity among rural agricultural populations, with multiple interconnected drivers contributing to this crisis [30]. According to other findings, [31] structural drivers, such as underdeveloped infrastructure, a lack of irrigation, and limited access to credit, perpetuate cycles of food insecurity in northern Ghana. Our findings reinforce this structural perspective, highlighting the need for systemic interventions rather than short-term, food-aid-based solutions.

The analysis revealed important nutritional consequences of food insecurity, particularly among mothers and children. Maternal haemoglobin levels were significantly lower in food-insecure households, indicating an increased risk of anaemia. Food insecurity was also associated with reduced household food expenditure, lower adherence to essential nutrition actions, and poorer child anthropometric outcomes, including height-for-age and weight-for-height Z-scores. The negative association between food insecurity and adherence to essential nutrition actions [ρ = - 0.213, p = 0.001] highlights how food scarcity undermines the adoption of recommended health and nutrition behaviours, thereby compounding nutritional risks.

The nutritional consequences identified in this study are consistent with international literature. Food insecurity has been widely linked to maternal anaemia, child stunting, and wasting in diverse low- and middle-income countries [LMIC] settings [32]. In our sample, the strongest correlation was observed between food insecurity and maternal haemoglobin levels [ρ = -0.283, p < 0.001], a result that has significant implications, given that anaemia contributes to maternal morbidity, adverse birth outcomes, and impaired productivity. The strong association between food insecurity and compromised child growth outcomes [height-for-age and weight-for-height] is also alarming, as stunting and wasting are not only immediate health risks but also predictors of reduced cognitive development, poorer educational attainment, and diminished economic productivity later in life [ 33; 34].

These findings align with several studies across multiple countries that demonstrate significant associations between household food insecurity and nutritional outcomes, particularly affecting mothers and children. Food insecurity increases the risk of maternal and child underweight [32]. It is strongly associated with an increased risk of anaemia, with a pooled analysis showing higher odds of anaemia [OR = 1.27] and iron-deficiency anaemia [OR = 1.45] among food-insecure populations [35]. Adult women and infants/toddlers in food-insecure households face particularly elevated anaemia risk [35]. Longitudinal studies reveal that food insecurity at age 5 significantly predicts lower height-for-age Z-scores across Ethiopia, India, Peru, and Vietnam, with effects persisting to age 8 [36]. In Cambodia, household food insecurity was positively associated with maternal thinness and anaemia, though child anthropometric outcomes showed less consistent associations [37]. The protective role of food security in both maternal and child nutrition outcomes suggests that interventions targeting household food security can generate intergenerational benefits. These findings stress the importance of integrating food security considerations into broader maternal and child health policies.

Maternal dietary quality was generally suboptimal in this study, with only 29.8% of women consuming all five essential food groups and an average dietary diversity score of 4.45 out of 10 possible groups. This finding aligns with national data from Ghana, where only 21% of adults consume all five food groups typically recommended in food-based dietary guidelines, including vegetables, fruits, pulses, nuts, and seeds, as well as animal-source foods and staple foods [38]. The national food group diversity score was 4.5 out of 10, which closely mirrors the study findings of 4.45 [38]. Furthermore, only 44% of women of reproductive age consume diets that meet minimum dietary diversity [MDD-W; at least 5 of 10 food groups], indicating that over half [56%] of women have a lower probability of micronutrient adequacy from their diet [38]. These findings are also consistent with previous research in northern Ghana, which reported similarly low levels of dietary diversity among women [39]. Low dietary diversity reflects both limited food availability and financial constraints, as well as entrenched social and cultural food practices [39;40].

Vegetable consumption was relatively adequate [76.2%], yet fruit intake was markedly lower [46.0%], producing an imbalance that restricts micronutrient adequacy. Inadequate fruit consumption is consistent with broader evidence from Ghana and other LMICs, where fruits remain less accessible and affordable compared to staple cereals and vegetables [42]. A study in Ghana [43;44] reports that vegetables were among the food groups most commonly consumed in addition to staple foods [87% of the population], while fruit was one of the food groups most likely to be missing from diets [46% of the population do not consume fruit]. The literature specifically identifies fruits as being consumed by less than half of the population, with consumption rates varying between urban dwellers [52%] and those in rural areas [37%] [43;44]. This dietary gap raises concerns about deficiencies in essential vitamins and minerals, particularly vitamin A, iron, and folate, which are critical for women of reproductive age.

The Global Diet Quality Score [Mean score of 11.97 out of 18] further indicated moderate overall diet quality. This finding is comparable to the national Global Dietary Recommendations [GDR] score of 10.5 out of 18 reported [44]. Literature [42] indicates that the Global Diet Quality Score [GDQS] assessment of women across three sub-Saharan African countries, including Ghana, showed poor overall diet quality, with 62-78% of women at moderate risk for nutrient inadequacy. However, the dietary transition toward increased consumption of energy-dense, nutrient-poor foods was evident, with 22.2% of women consuming sweet foods and 21.8% reporting multiple sugary sweetened beverages. In Ghana, it is reported that [44], sweet foods are consumed by 40% of urban populations versus 27% of rural populations, while soft drinks are consumed by 21% of urban populations versus 12% of rural populations. The literature emphasises that these foods are known to displace nutrient-rich foods in the diet and are associated with an elevated risk of NCDs [Ghana Diet Quality Profile, 2022]. These patterns align with reports of rising sugar consumption and the gradual nutrition transition in Ghana [43;44;45]. This dual burden of persistent undernutrition coupled with growing exposure to unhealthy diets underscores the complexity of nutrition challenges facing Ghanaian women, particularly in rural areas.

This study highlights a paradox in infant and young child feeding, where breastfeeding practices are exemplary, yet complementary feeding is markedly inadequate. Almost all children [97.2%] had ever been breastfed, and 90.6% continued breastfeeding into the second year of life, aligning with cultural traditions in Ghana and echoing previous reports of breastfeeding rates ranging from 92% to 97% [46]. In contrast, complementary feeding indicators were suboptimal, with only 44.4% achieving the minimum dietary diversity [MDD], 51.5% meeting the minimum meal frequency [MMF], and 27.2% achieving the minimum acceptable diet [MAD]. Although these figures are somewhat higher than national averages, MDD 25.46%, MMF 32.82%, and MAD 11.72% [47], they remain well below global recommendations and are consistent with regional studies reporting MDD between 10.5% and 51.4% and MAD between 8.5% and 29.9% [46; 48]. The disparity between breastfeeding and complementary feeding outcomes reflects structural and knowledge-related challenges: while breastfeeding is strongly supported by cultural norms, adequate complementary feeding requires access to diverse foods, time, and maternal knowledge of age-appropriate practices. Determinants such as maternal education, household wealth, urban residence, and maternal knowledge have been repeatedly identified as key influences [47; 46; 48]. Thus, despite relatively better performance in the study area compared with national averages, complementary feeding deficiencies persist and threaten child growth and development, underscoring the urgent need for interventions that combine caregiver education with strategies to improve access to diverse, affordable, and nutrient-rich foods.

Another concerning finding was the high consumption of unhealthy foods among children, with 41.6% consuming sugar-sweetened beverages and 25.0% consuming sweet foods. These findings align with regional patterns where Becher *et al.* [49] documented similar consumption rates among northern Ghanaian children [46% sugar-sweetened beverages, 51% salty snacks], and Rousham *et al.*, [45] found 39.9% consuming sugar-sweetened beverages across Ghana and Kenya. Early introduction of energy-dense, nutrient-poor foods establishes unhealthy dietary preferences and increases long-term obesity risk [53].

The presence of such patterns in rural northern Ghana underscores the pervasiveness of the nutrition transition since the 1970s [53], even in resource-constrained settings. This reflects broader West African trends where children consume energy-dense foods seven times more frequently than fruits and vegetables, with adult obesity increasing 115% over 15 years [54].

The associations of food security and dietary quality identified in this study were household food expenditure, household structure [polygamy versus monogamy], and food security status itself.

These associations align with established research demonstrating that household economic capacity serves as the primary driver of nutritional outcomes across diverse contexts. Monthly food expenditure emerged as the strongest predictor of dietary diversity [Z=-6.558, p<0.001, r=-0.44], confirming the central role of household economic resources in determining food access and quality. This finding is consistent with research from rural Ethiopia, where higher food expenditure demonstrated the strongest association with dietary diversity among households [55]. Similarly, our results showing that children from families with higher food expenditure achieved significantly better dietary diversity [Z=-6.260, p<0.001, r=-0.397] mirror findings from Bangladesh, where children from food-secure households exhibited 26% higher dietary diversity scores, with household wealth showing strong positive associations with dietary outcomes [56].

Poorer dietary quality was observed among women in polygamous households, compared to those in monogamous unions. Children from monogamous households achieved significantly better dietary diversity compared to those in polygamous households [p=0.003]. This trend may be explained by resource dilution, whereby larger household sizes and competing demands limit women’s and children’s access to diverse foods. Earlier research reports that in northern Ghana, children of monogamous mothers demonstrate better stature than children of second wives [57]. Women in monogamous households achieve significantly higher dietary diversity compared to those in polygamous unions, with monogamous women having 1.42 times higher odds of achieving better dietary diversity [58]. However, findings from Nigeria present contrasting evidence, showing polygynous households have better food security at the household level, though children of polygynous mothers experience worse long-term nutrition outcomes [59]. Addressing the nutritional vulnerability of women in polygamous households, therefore, requires culturally sensitive interventions that recognise household resource allocation dynamics.

An additional critical contributing factor was health insurance coverage, with uninsured households reporting significantly higher food insecurity rates [84.4%] compared to those with insurance [72.6%]. These findings align with previous research that demonstrates the interconnectedness of healthcare access and food security. Antabe *et al.*, [2019] found that in Ghana, severely food-insecure households were significantly less likely to enrol in national health insurance compared to food-secure households [OR = 0.36, P < .05], illustrating how food insecurity can create barriers to accessing healthcare.

Himmelstein, [2019] documented that Medicaid expansion under the Affordable Care Act was associated with a significant 2.2-percentage-point decline in very low food security rates among low-income adults, representing a 12.5% relative reduction. These finding highlights the interconnection between healthcare access and food security. By reducing out-of-pocket healthcare costs, health insurance appears to protect household resources that would otherwise be diverted from food expenditure.

Partner occupation also influenced household food security. Households in which partners were engaged in trading reported significantly lower food insecurity [37.5%] compared to farming households [82.4%]. This disparity highlights the heightened vulnerability of subsistence farmers to seasonal production variability, market fluctuations, and climate shocks, while traders benefit from more stable and diversified income streams that enhance food access. These findings align with evidence from Nigeria, where households engaged in off-farm activities experience better food security outcomes than those relying solely on crop production, with diversified households recording surplus indices of 0.71 compared to 0.62 for crop-only households [62]. Similarly, trading activities have been shown to provide steadier income sources than farming, thereby reducing susceptibility to seasonal and climatic shocks[63]. More broadly, livelihood diversification is recognised as a key resilience strategy, with non-agricultural activities such as trading and manufacturing contributing more directly to income stability than agricultural diversification alone, although effectiveness is mediated by access to resources and supportive local infrastructure [64].

The strikingly high prevalence of maternal illiteracy [84.3%] represents a critical socio-demographic factor shaping nutrition outcomes in this context. Education is strongly linked to health and nutrition knowledge, household decision-making, and income-earning opportunities [65], making the low educational attainment among women in this study particularly concerning. Mensah-Bonsu [2023] also recommends improved education, social structures, and support services to combat food insecurity. This limited education likely constrains awareness of optimal dietary practices and reduces women’s ability to negotiate household resource allocation. These educational barriers are further reinforced by research from Zakaria [38], who found that literacy was a significant socioeconomic determinant of dietary diversity among women of childbearing age, with over half having inadequate dietary diversity scores below 5. When combined with economic constraints and gendered household structures, low maternal education compounds the multiple barriers to achieving adequate diet quality in rural northern Ghana.

This study’s findings reinforce the need for poverty alleviation and women’s economic empowerment as central strategies for improving maternal and child nutrition. Income-generating interventions and skills development initiatives targeting women can directly enhance food access and dietary diversity. Also, the protective effect of health insurance coverage against food insecurity suggests that policies promoting universal health coverage can yield dual benefits, improve healthcare access while also safeguarding household resources for food. This highlights the potential for integrated social protection policies that simultaneously address health and nutrition.

Furthermore, the negative impact of polygamous households on dietary quality underscores the importance of considering cultural and household structures in nutrition programming. While respecting cultural norms, interventions should explicitly address intra-household resource distribution and provide targeted support to women and children in polygamous households. the nutrition patterns also observed, particularly increased consumption of sugary foods and beverages among both mothers and children, signal the need for proactive prevention of non-communicable diseases. Nutrition education campaigns should not only promote dietary diversity but also raise awareness about the health risks of processed and sugary foods, even in rural contexts where undernutrition remains prevalent. The interconnected nature of maternal and child nutrition outcomes emphasises the importance of codesigned and context and culturally specific interventions to address intergenerational nutritional challenges effectively in resource-poor settings.

### Study Strengths and Limitations

This study provides one of the few comprehensive assessments of both maternal and child dietary adequacy alongside food security in rural northern Ghana. However, the cross-sectional design limits the ability to establish causality between food insecurity, dietary quality, and nutritional outcomes. Self-reported dietary data and household food security measures may be subject to recall bias, potentially affecting accuracy. The study’s focus on a single municipality also limits generalizability to other regions of Ghana or similar populations elsewhere in sub-Saharan Africa. There is also the potential for social desirability bias, where participants may underreport food insecurity or overreport healthy food consumption. Future research should adopt longitudinal designs to capture seasonal and temporal variations in food insecurity and nutrition outcomes and explore causal pathways more rigorously.

### Conclusions

This study demonstrates that household food insecurity remains pervasive in the East Mamprusi Municipality, with profound consequences for maternal and child nutrition. Dietary inadequacy among women and young children reflects the combined effects of economic capacity, household structure, and access to resources. While breastfeeding practices remain strong, complementary feeding is insufficient, and unhealthy dietary patterns are emerging even in resource-constrained settings.

Interventions should prioritise women’s economic empowerment, universal health coverage, and culturally sensitive approaches that address household dynamics such as polygamy. Future initiatives could focus on promoting climate-resilient and nutrition-sensitive agriculture, improving food storage and market access to mitigate seasonal food insecurity, and evaluating the effectiveness of integrated livelihood and nutrition programmes. Further research should explore how seasonal food availability and intra-household decision-making influence dietary diversity and long-term nutritional outcomes.

## Declarations

### Ethics approval and consent to participate

The study received ethical approval from the Committee on Human Research, Publications, and Ethics at Kwame Nkrumah University of Science and Technology **[CHRPE/AP/149/23]** following permission from the Ghana Health Service and the East Mamprusi Municipal Health Directorate **[GHS/NER/EMM/10/2023].** Confidentiality and anonymity were maintained throughout the study to protect participants’ privacy. Each participant was assigned a unique identification code instead of using personal identifiers such as names or addresses. Completed questionnaires were securely stored, accessible only to the research team. Reports are presented in aggregated form, ensuring that individual participants cannot be identified. In addition, verbal and written informed consent were obtained before participation, with a clear explanation that participants could withdraw at any time without any consequences.

### Consent for publication

Not applicable

### Availability of data and materials

Researchers may request the corresponding author for access to the datasets used and/or analysed in this study.

### Authors’ contributions

Reginald A. Annan [RAA] and Charles Apprey [CA] are affiliated with the Department of Biochemistry and Biotechnology, Kwame Nkrumah University of Science and Technology, Kumasi, Ghana. Vincent Adocta Awuuh [VAA] is associated with both the Department of Biochemistry and Biotechnology at Kwame Nkrumah University of Science and Technology, Kumasi, Ghana, as a student and the Department of Nutrition and Dietetics, College of Nursing and Midwifery, Nalerigu, Ministry of Health, Ghana. VAA, RAA, and CA contributed substantially to the conception and design of the study, data acquisition, and interpretation of findings. VAA performed the data analysis and drafted the initial version of the manuscript. RAA and CA critically reviewed and revised the manuscript for important intellectual content. All authors approved the final version of the manuscript and agree to be accountable for all aspects of the work.

## Data Availability

Data is available upon request to the researchers

## Acknowledgements

We sincerely thank all participants in this study and extend our best wishes to them. We also acknowledge the Canadian Queen Elizabeth II Diamond Jubilee Advanced Scholars – West Africa: Nutrition Research Capacity Building, Netlinks for Enhanced Health Equity and Sustainable Inclusive Growth in Rural West Africa, for their invaluable contribution to capacity building during this PhD journey. Special recognition is given to the Support Programme for Postgraduate Students, North-West University, Potchefstroom, South Africa, for their continuous support and commitment to academic and research empowerment. Finally, our heartfelt gratitude goes to the College of Nursing and Midwifery, Nalerigu, for their unwavering logistical support and dedicated assistance throughout data collection and laboratory analysis, which significantly contributed to the successful completion of this study.

